# Classification of Variants of Reduced Penetrance in High Penetrance Cancer Susceptibility Genes: Framework for Genetics Clinicians and Clinical Scientists by CanVIG-UK (Cancer Variant Interpretation Group-UK)

**DOI:** 10.1101/2024.07.19.24310706

**Authors:** Alice Garrett, Sophie Allen, Miranda Durkie, George J Burghel, Rachel Robinson, Alison Callaway, Joanne Field, Bethan Frugtniet, Sheila Palmer-Smith, Jonathan Grant, Judith Pagan, Trudi McDevitt, Charlie F. Rowlands, Terri McVeigh, Helen Hanson, Clare Turnbull, CanVIG-UK

**Author notes:** Corresponding author: Prof. Clare Turnbull. these authors contributed equally to this manuscript (Alice Garrett, Sophie Allen).

## Abstract

**Purpose:** Current practice is to report and manage likely pathogenic/pathogenic variants in a given cancer susceptibility gene (CSG) as though having equivalent penetrance, despite increasing evidence of inter-variant variability in risk associations. Using existing variant interpretation approaches, largely based on full-penetrance models, variants where reduced penetrance is suspected may be classified inconsistently and/or as variants of uncertain significance (VUS). We aimed to develop a national consensus approach for such variants within the Cancer Variant Interpretation Group UK (CanVIG-UK) multidisciplinary network.

**Methods:** A series of surveys and live polls were conducted during and between CanVIG-UK monthly meetings on various scenarios potentially indicating reduced penetrance. These informed the iterative development of a framework for the classification of variants of reduced penetrance by the CanVIG-UK Steering and Advisory Group (CStAG) working group.

**Results:** CanVIG-UK recommendations for amendment of the 2015 ACMG/AMP variant interpretation framework were developed for variants where (i) Active evidence suggests a reduced penetrance effect size (e.g. from case-control or segregation data) (ii) Reduced penetrance effect is inferred from weaker/potentially-inconsistent observed data.

**Conclusions:** CanVIG-UK propose a framework for the classification of variants of reduced penetrance in high-penetrance genes. These principles, whilst developed for CSGs, are potentially applicable to other clinical contexts.

## Introduction

### Introduction to Variant Interpretation

With rapid advances in high-throughput molecular and bioinformatic pipelines for genomic sequencing, the bottleneck in genetic testing has largely moved to the clinical interpretation of detected variants. With a historical landscape of disparate classification methodologies and conflicting interpretations, the improved consistency afforded via near global adoption of the 2015 ACMG/AMP framework from Richards et al has been transformative^1^. The original 2015 ACMG/AMP framework has been augmented by (i) transformation in 2018 by the ClinGen Sequence Variant Interpretation (SVI) group from the original “categorical” approach into a more quantitative Bayesian system of evidence quantitation and (ii) detailed specification of the framework for selected gene-phenotype dyads by ClinGen Variant Curation Expert Panels (VCEPs).^2–5^ Supporting, moderate, strong and very strong evidence from the original ACMG/AMP framework are assigned 1, 2, 4 and 8 “evidence points” respectively within the 2018 ClinGen Bayesian evidence quantitation system. Overall classifications would be assigned based on the net evidence points: pathogenic (≥10), likely pathogenic (6-9), likely benign ((−1)-(−5)) and benign (≤-6)^3^ ^4^

### Current clinical paradigms of disease penetrance

There is *a priori* biological likelihood and observed evidence for different variants (ascribed as pathogenic within the same gene) having different risks of disease. Missense variants might be anticipated to have different clinical impacts relating to differing effects on protein conformation and binding; variants altering splicing might be anticipated to have different clinical impacts according to the consequent proportions of isoforms. Furthermore, variability in penetrance would not necessarily be proportionate for the differing constituent phenotypes associated with a gene. For example, analyses demonstrate protein-truncating variants (PTVs) in different regions of the *BRCA1* and *BRCA2* genes exhibit differential risks of ovarian cancer versus breast cancer^6^ ^7^. For *BRCA1*, missense pathogenic variants in the RING domain exhibit lower risks than PTVs, whilst for *TP53,* missense pathogenic variants in the DNA-binding domain exhibit higher risks than PTVs^8^ ^9^. However, all of these observations are derived from collapsed analyses of groups of eligible variants; examples robustly demonstrating reduced penetrance for individual variants are very rare. Through diligent collaborative international assembly of multi-case families, the ENIGMA consortium demonstrated via segregation analyses that the *BRCA1* variant NM_007294.4:c.5096G>A p.(Arg1699Gln) is associated with risks of breast and ovarian cancer that are statistically significantly lower than the risks for standard *BRCA1* pathogenic variants^10^ ^11^.

### Variant classification where data suggest reduced penetrance

The most straightforward evidence of reduced penetrance is a directly quantified measure of disease association, typically an odds ratio from comparison of unselected cases against controls (but potentially also from a segregation/family history analysis performed under a reduced penetrance model). Ancillary evidence suggestive of and consistent with reduced penetrance may include: (i) Evidence towards pathogenicity of insufficient strength to gain evidence points in the context of full penetrance, hereafter termed “*weakly-pathogenic*” evidence (e.g. intermediate/conflicting results from functional assays) and (ii) Evidence that would support benignity in the context of full penetrance, hereafter termed “*potentially-contradictory*” evidence (e.g. biallelic case with no/minimal relevant phenotype). Another “*potentially-contradictory*” evidence type would be apparent non-segregation of the variant with disease (analysed under a high penetrance model); a variant of reduced penetrance would yield weaker segregation as well as there being a high phenocopy rate for common diseases such a breast cancer.

We present here consultative development of a consensus framework for the classification of variants of reduced penetrance. We used as our reference and source of exemplars, hereditary breast and ovarian cancer (HBOC) genes *BRCA1/BRCA2*, for which (i) reduced penetrance is frequently raised clinically, (ii) multiple sources of case-control, clinical and functional data are available and (iii) definitions for breast cancer susceptibility genes are already established for “high-penetrance” (OR>4) and “moderate-penetrance” (OR 2-4)^12^. We considered how evidence items might be incorporated in a classification of “Likely pathogenic-reduced penetrance” addressing two distinct scenarios:

**Scenario A:** in which there is active quantitative evidence of an effect size consistent with reduced penetrance (e.g. breast cancer OR 2-4)^12^

**Scenario B**: in which there is no active quantitative evidence of effect size; several evidence items indicate pathogenicity but at least one piece of contributary evidence would be considered *potentially-contradictory* or would be too *weakly-pathogenic* in a standard penetrance context and could indicate pathogenicity with reduced penetrance.

## Methods

### Overview

The consensus framework (Figure 1) was iteratively developed between September 2023 and July 2024 (Table 1), leveraging the existing organisational structures of:

i. **Cancer Variant Interpretation Group UK (CanVIG-UK) (Supplementary Note 1):** a national group comprising >300 clinical scientists, clinical geneticists and genetic counsellors from the UK and the Republic of Ireland^13^. CanVIG-UK was convened in 2017 at the request of the UK Association for Genomic Science (ACGS), to advance UK consistency in application of the 2015 ACMG/AMP framework for variant interpretation in Cancer Susceptibility Genes (CSGs)^14^. CanVIG-UK holds monthly meetings (of 1 hour 15 minutes) at which issues related to variant interpretation and challenging cases are discussed to reach national consensus. Leveraging CanVIG-UK structures, comprising email circulations and monthly meetings, we conducted surveys and polls to understand views and practice from the UK clinical-diagnostic community.
ii. **The CanVIG-UK Steering and Advisory Group (CStAG) (Supplementary Note 2):** CStAG comprising 13 expert clinical scientists and clinical geneticists, providing oversight and strategy for CanVIG-UK. CStAG holds separate monthly meetings (1 hour 45 minutes). CStAG served as a working group for drafting of the framework and development of the case-examples to be used with CanVIG-UK.

**Figure 1:**
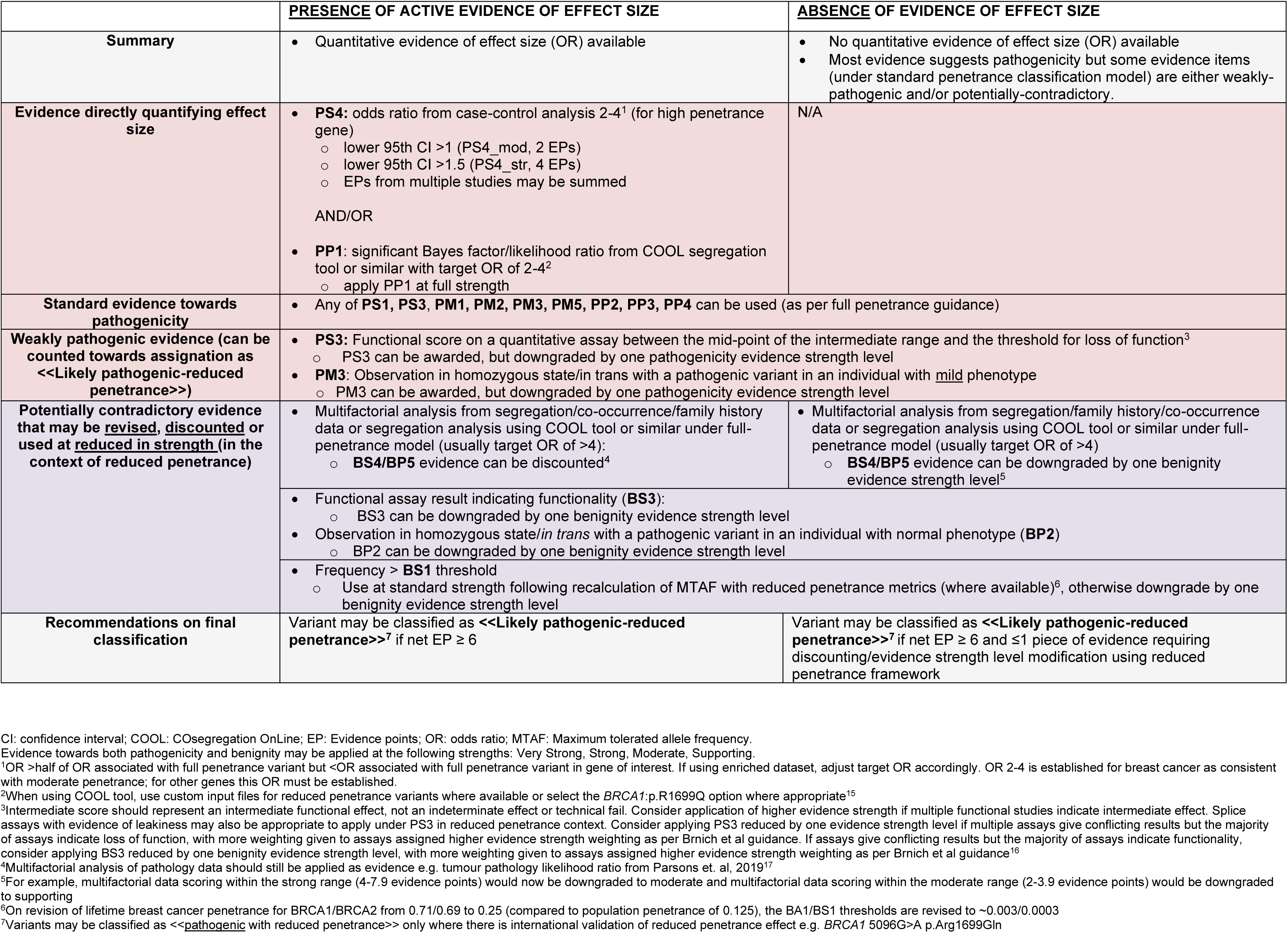
Consensus framework developed iteratively after consultation with CStAG and the CanVIG-UK Clinical Network. CI: confidence interval; COOL: COsegregation OnLine; EP: Evidence points; OR: odds ratio; MTAF: Maximum tolerated allele frequency. Evidence towards both pathogenicity and benignity may be applied at the following strengths: Very Strong, Strong, Moderate, Supporting.

**Table 1:** Stages in CanVIG-UK consultative development of framework for the classification of variants of reduced penetrance. (see Supplementary Methods for further details). Following feedback at stages 3-4, the initial draft framework was adjusted by the CStAG drafting group and re-circulated.

|  | Date | Stage | Format | Brief Description |
| --- | --- | --- | --- | --- |
| 1 | Aug-Sep 2023 | Scoping survey | Online Survey (Email send-out) | Initial survey on existing practice and confidence in classifying variants of reduced penetrance |
| 2 | Sep 2023 | Evidence Allocation poll | Live poll | Poll to explore how evidence might be allocated (hypothetical variant scenarios) |
| Initial draft framework created and circulated |  |  |  |  |
| 3 | Feb 2024 | Variant Scenario A: Active quantitative evidence of effect size | Live poll | Poll to test framework application (real exemplar variant scenarios) |
| 4 | Feb-Mar 2024 | Variant Scenario B: No active quantitative evidence of effect size | Online Survey (Email send-out) and Live poll | Poll and survey to test framework application (real exemplar and hypothetical variant scenarios) |
| Pre-final framework created and circulated |  |  |  |  |
| 5 | July 2024 | Finalisation | Live CanVIG-UK presentation | Final amendments following peer review |
| Final framework created and circulated |  |  |  |  |

## Results

### Responses to Scoping Survey

For the scoping survey, there were 37 respondents representing 19 different genetics services (covering all 7 English genomic laboratory hubs). 20/37 (54%) respondents were clinical scientists, 3/37 (8%) were trainee clinical scientists, 7/37 (19%) were consultant clinical geneticists, and 7/37 (19%) had other roles (genetic counsellors, molecular pathologists and oncologists). Over half of respondents (19/37, 51%) reported that their laboratory service had newly classified a variant as being of reduced penetrance locally without recourse to national discussion. The frequency of local discussions regarding potential for a CSG variant being of reduced penetrance varied widely, with 2/37 (0.5%) respondents reporting frequency being weekly and 10/37 (27%) reporting a frequency of six-monthly (or less often); the most common frequency was every two to six months (13/37, 35%) (see Supplementary Figure 1). Respondents’ confidence in various contexts of classifying variants as being of reduced penetrance are shown in Figure 2, with confidence generally reported as lower for classifications undertaken as an individual compared to after a local multidisciplinary team (MDT) discussion compared to discussion at national level. 37/37 respondents believed that it would be “extremely” or “very” valuable” for CanVIG-UK to develop a framework for reporting variants of reduced penetrance.

**Figure 2:**
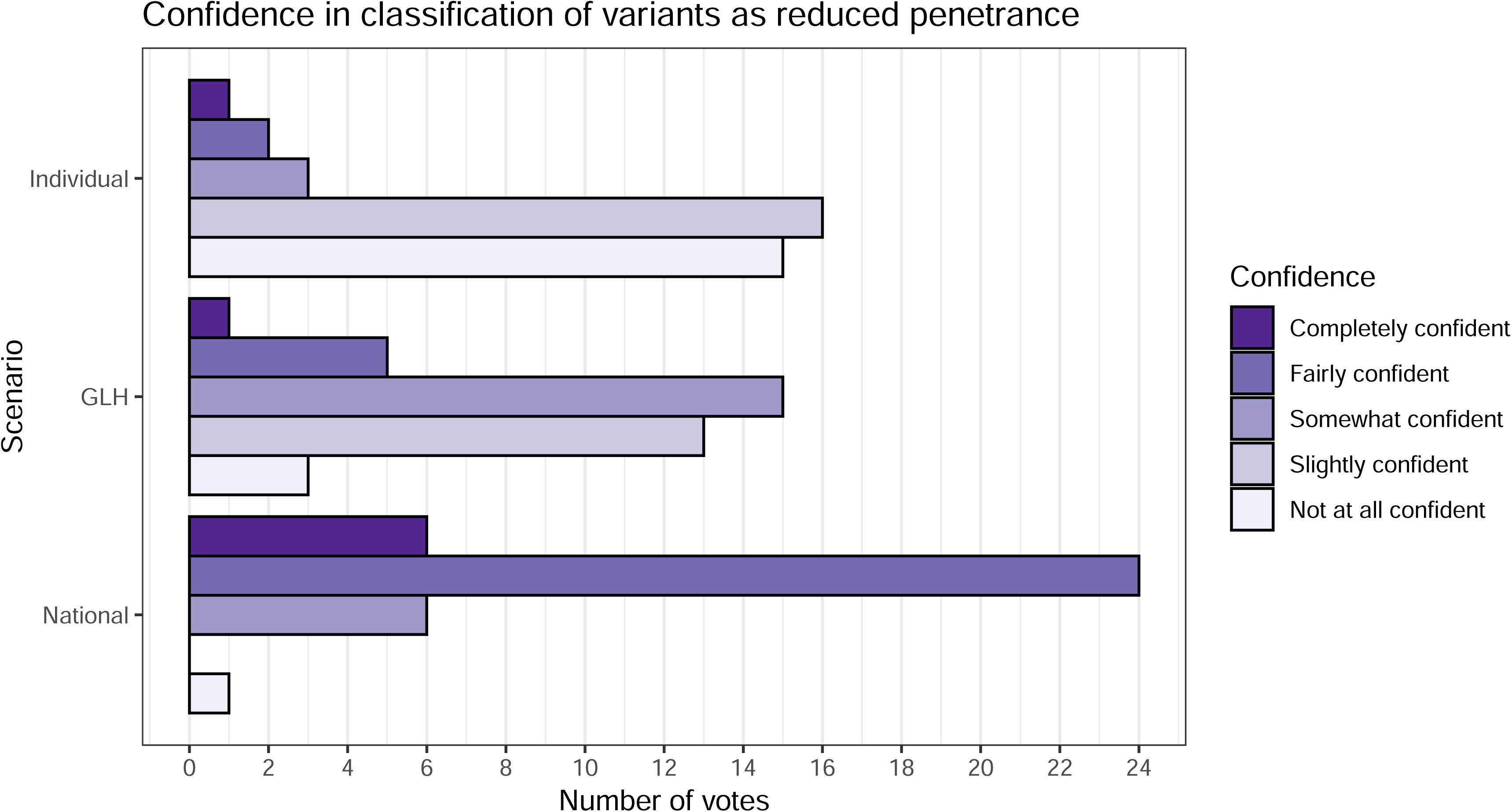
Confidence of survey respondents in classifying variants as reduced penetrance. Respondents ranked their confidence on a scale from 1-5 (1=Not at all confident, 5=Completely confident) in three scenarios: a) as an individual without group discussion (Individual), b) after discussion at a local review meeting (Genomic Laboratory Hub (GLH) review), and c) after discussion at a national-level meeting (National).

### Responses to Evidence Allocation Poll

26/66 (39%) attendees at the September 2023 CanVIG-UK monthly meeting participated in the in-meeting evidence allocation poll: 17/26 (65%) clinical scientists, 5/26 (19%) trainee clinical scientists, 3/26 (12%) clinical genetics consultants and 1/26 (4%) clinical genetics trainee. In this poll, we explored for hypothetical variants with existing evidence towards pathogenicity how participants would allocate evidence points for various types of observations for: (i) PS4 (case-control data), (ii) PS3/BS3 (functional assay data), (iii) BP2/BS2/PM3 (observation of a variant in trans) and (iv) BS4 (segregation data) if being classified under a framework of reduced penetrance (see Supplementary Table 1).

Under a reduced penetrance classification framework, the majority of participants endorsed application of case-control evidence (PS4) at supporting or moderate level if point estimate of OR>2 and lower 95% CI>1 (17/23, 74% endorsed) and up to strong evidence where OR>2 and lower 95% CI>1.5 (25/26, 96% endorsed). For functional evidence that might be ascribed as “*weakly-pathogenic*”, for example where a quantifiable functional assay yielded a result in the upper intermediate range (towards the loss of function), most participants (18/22, 82%) elected to apply PS3 but at a reduced strength. When asked how they would consider “*potentially-contradictory*” evidence items, for example (i) data against segregation that was generated under a full penetrance model or (ii) observation of the variant *in trans* with a known pathogenic variant in the absence of manifest phenotype, that majority of participants (13/23, 57%) down-scored the evidence points towards benignity if the variant was to be assigned as ‘reduced penetrance’.

### Responses to “Scenario A” Variant Classification Polls

Having pre-circulated a draft Reduced Penetrance Framework (Supplementary Figure 2), we undertook live polling to explore application of the framework for real variants for which active quantitative evidence of effect size are available (“Scenario A” variants). The evidence items for each variant were presented live and participants were invited to provide a final classification based on these evidence items (summarised in Table 2). A classification of “Likely pathogenic-reduced penetrance” was asserted by 76% of respondents for the first variant (*BRCA2* NM_000059.4:c.9302T>G p.(Leu3101Arg)) and by 81% for the second variant (*BRCA2* NM_000059.4:c.520C>T p.(Arg174Cys)) (Figure 3A, 3B). When asked by live poll if they were confident using the framework for classifying a variant as being of reduced penetrance where there is a quantified measure of effect size (that is, “Scenario A”), there was an overall positive response (25/27, 93% and 21/27, 78%, of respondents agreed they were confident where the quantified effect size came from case-control data or segregation data respectively) (Supplementary Table 2).

**Figure 3:**
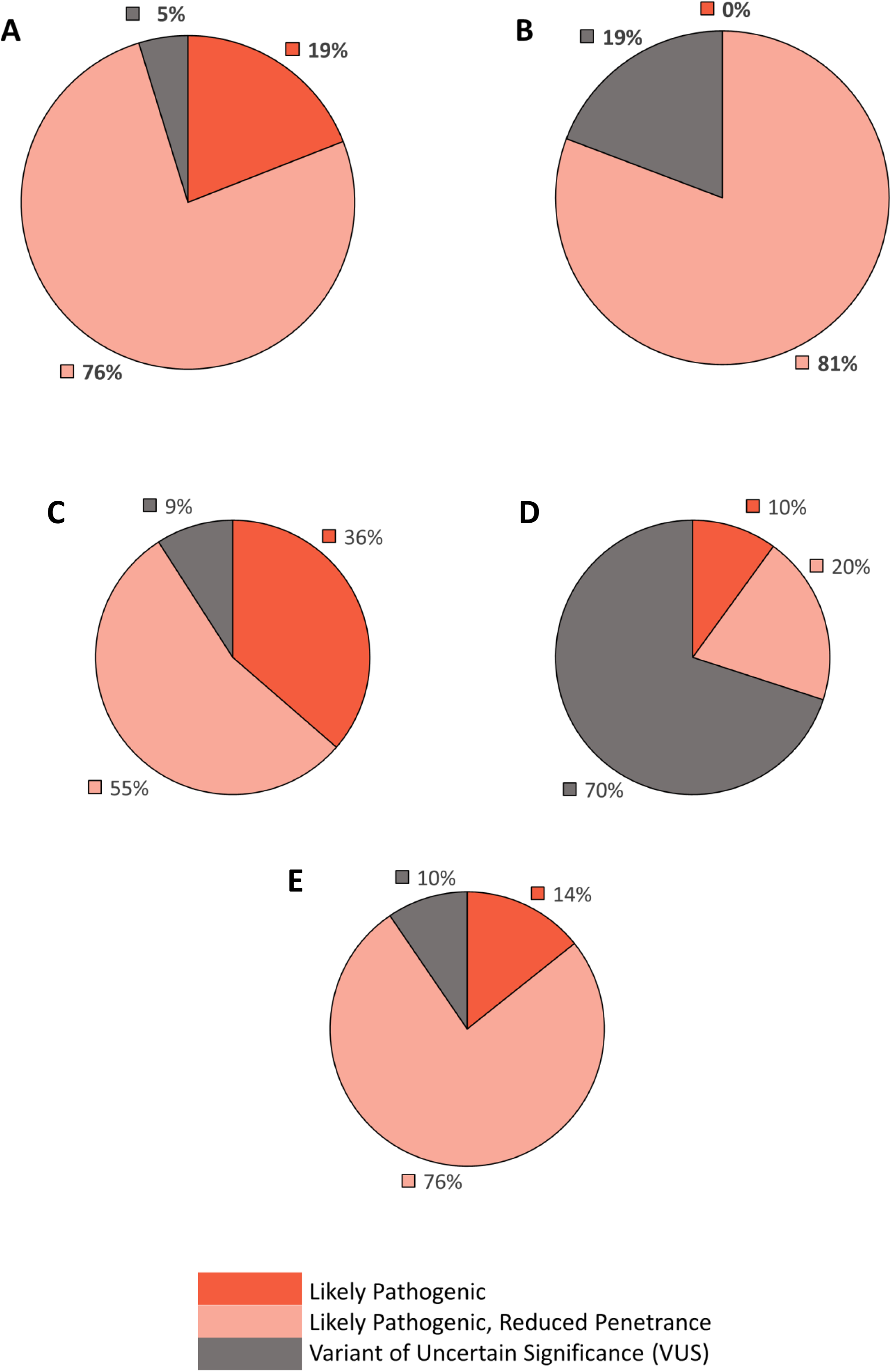
CanVIG-UK Poll results for classification of two *BRCA2* Scenario A variants and three hypothetical Scenario B variants using the framework. See Table 2 for evidence items applied for these variant scenarios. A: Classifications for *BRCA2* NM_000059.4:c.9302T>G c.9302T>G p.(Leu3101Arg). B: Classifications for NM_000059.4:c.520C>T p.(Arg174Cys). C: Hypothetical variant scenarios. Variant_i: substantial evidence towards pathogenicity but normal homozygote reported in literature. Variant_ii: some evidence towards pathogenicity but segregation/family history data towards benignity under a full penetrance model. Variant_iii: substantial evidence towards pathogenicity but segregation/family history data towards benignity under a full penetrance model.

**Table 2:**
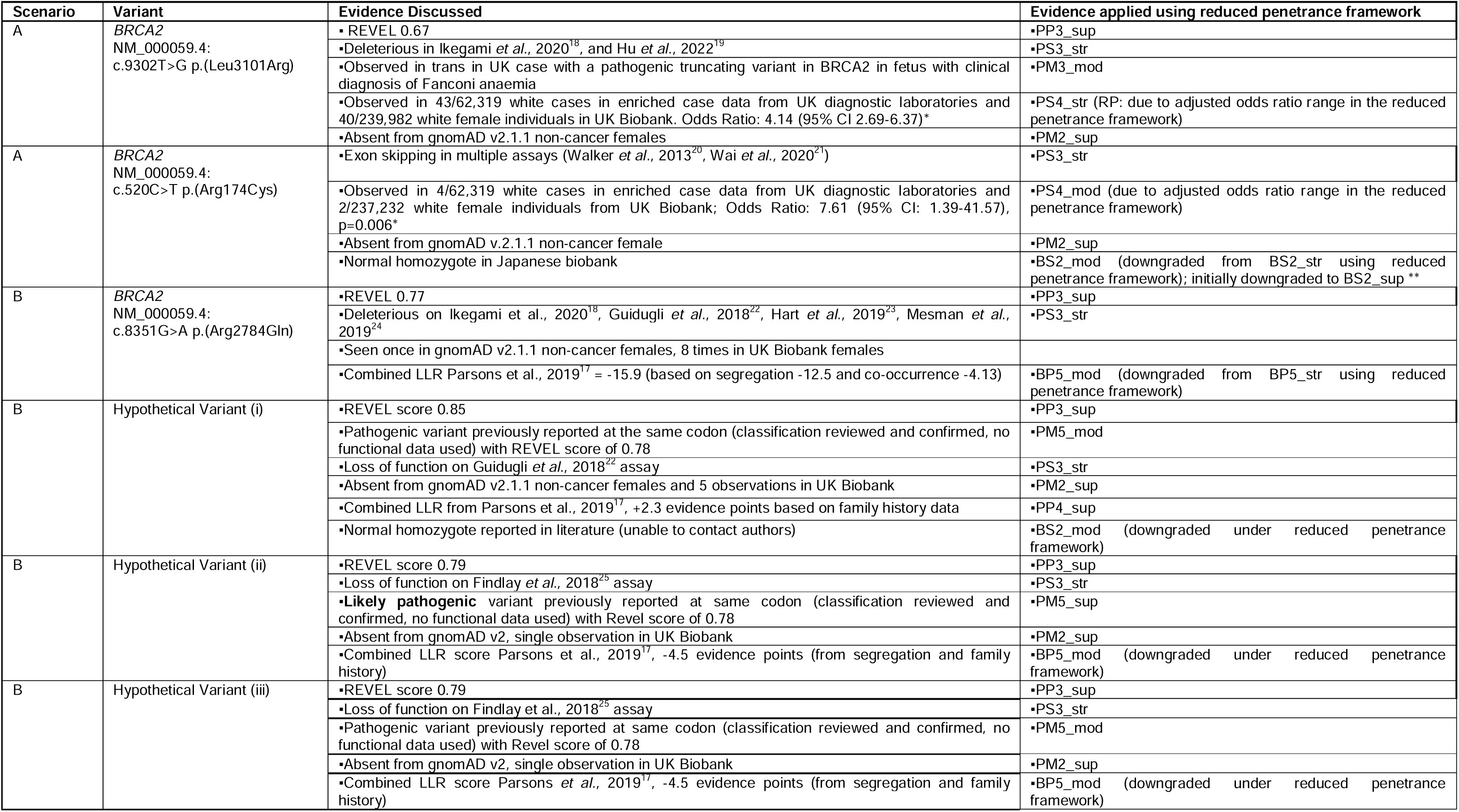
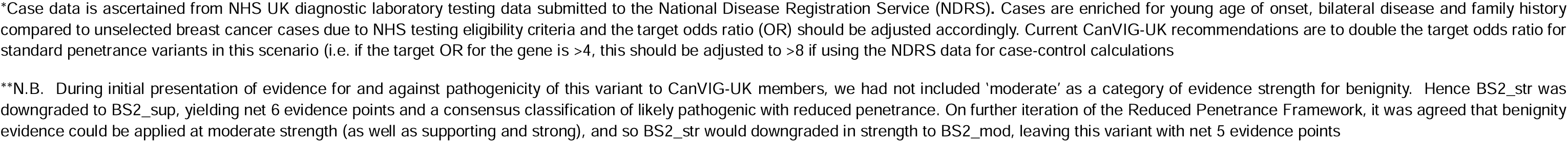
Exemplar and hypothetical variants discussed and voted on by live poll in CanVIG-UK meetings (February-March 2024) during development of the framework.

### Responses to “Scenario B” Variant Classification Polls

The framework for addressing “Scenario B” variants was evolved over two live meetings in consecutive months (February and March 2024) with intervening surveys and amendment to the framework. Following a live poll regarding classification of a real “Scenario B” variant (*BRCA2* NM_000059.4:c.8351G>A p.(Arg2784Gln) (Supplementary Figure 3)), only 7/26 (27%) respondents reported feeling comfortable using the framework for scenario B variants (Supplementary Table 2). During subsequent live discussion, concerns were articulated that the framework could allow too much use of “*potentially-contradictory”* or only “*weakly-pathogenic”* evidence. In response to this feedback, CStAG undertook review and amendment to the framework to restrict the number of such evidence items applicable. We then tested response to these revisions through creation of hypothetical variants for “Scenario B” (featuring a range of evidence items for scoring, summarised in Table 2).

There was majority agreement (Figure 3) around how these hypothetical variants would be classified both from responses via the online survey for Variant_i (6/11 (55%) classified as Likely Pathogenic-reduced penetrance) and for Variant_ii (7/10 (70%) classified as variant of uncertain significance (VUS)) and for a third variant classified via live polling (Variant (iii): 16/21 (76%) classified as Likely Pathogenic-reduced penetrance). Following live classification of Variant_iii, in discussion and subsequent live poll 15/24 (63%) of CanVIG-UK respondents reported feeling confident in applying the revised framework for “Scenario B” variants (Supplementary Table 2).

### Reduced penetrance variant classification framework

Figure 1 shows the CanVIG-UK modified variant classification framework for reduced penetrance variants in high penetrance cancer susceptibility genes: in Scenario A there is active quantified evidence of an effect size (left half of framework) whilst Scenario B covers variants for which such data are not available (right half of framework). Recommended evidence applications and adjustments vary between these scenarios on account of the prior level of confidence in effect size. Scoring adjustments for potentially-contradictory or weakly-pathogenic evidence for the two scenarios are delineated.

## Discussion

We present here a framework for classification of variants of presumed reduced penetrance observed in high-penetrance cancer susceptibility genes. This framework was developed and tested within the CanVIG-UK community of diagnostic clinical scientists, clinical geneticists and other healthcare professionals working in cancer susceptibility genetics. Via an initial scoping survey, we identified substantial heterogeneity in practice in regard of local stand-alone classification of CSG variants as being reduced penetrance. In a live evidence allocation poll (using synthetic scenarios for which other concomitant evidence items towards pathogenicity were pre-stated), there was again substantial variation between participants in the evidence points they proposed for allocation under a reduced-penetrance-type scenario for PS4 (case-control data), PS3/BS3 (functional assay data), BP2/BS2/PM3 (observation of a variant in trans) and BS4 (segregation data). This indicated the potential value of harmonising practice via a consensus framework. Via the CStAG working group, we developed a framework for classification of variants of reduced penetrance covering two distinct use-cases. We defined as “Scenario A” variants with available active quantitative evidence of an effect size consistent with reduced penetrance (e.g. OR 2-4 for breast cancer for *BRCA1/BRCA2* variants)^12^. We defined as “Scenario B” variants where there was no active quantitation of effect size, but sizeable evidence indicating pathogenicity for which some items were only *“weakly-pathogenic”* and/or there was evidence which was “*potentially-contradictory*” (evidence counting towards benignity when scored under a model of full penetrance). Application of the framework was tested and iterated with the CanVIG-UK community using live polls and email send-out surveys comprising both real and hypothetical variants under each of Scenario A and B. Following in-meeting live discussion, the framework was refined to stipulate (i) that for Scenario B, no more than one *“weakly-pathogenic”* or “*potentially-contradictory*” evidence item can be included, (ii) “Likely pathogenic-reduced penetrance” as the only resultant classification (in the absence of preceding *international* consensus e.g. the established *BRCA1* reduced penetrance *BRCA1* variant NM_007294.4:c.5096G>A p.(Arg1699Gln) may be classified as pathogenic with reduced penetrance).

### Application of Framework in other clinical gene-disease paradigms

Whilst the variant examples considered were for the high-penetrance CSGs *BRCA1/BRCA2*, the framework could be applied in other disease areas, for example inherited colorectal cancer or cardiac disease, for which case-control data exist and providing effect-sizes constituting high- and moderate-penetrance have been established.

The magnitude of effect size (i.e. relative risk/odds ratio for association) ascribed as reduced penetrance is highly context-specific. For a typical rare monogenic disease, the lifetime risk might be 1 in 10,000, meaning that a variant of standard (high) penetrance is potentially conferring a relative risk (or odds ratio) of several thousand-fold. In this context, there is plenty of “risk space” residing between that effect size for standard (high) penetrance and neutrality (OR=1). Indeed in 2023, ClinGen delineated a model of three zones of risk (high-penetrance, low penetrance, risk allele) that would fit the typical rare monogenic disease paradigm, and considered in this context how “risk alleles” should be classified and managed^26^. For breast cancer, the female population lifetime absolute risk is 1 in 8 (12.5%) and a variant is deemed as high-penetrance if the OR≥4; this leaves a relatively tight “risk-space” in which to delineate a zone of reduced penetrance. Nevertheless, there is consistency in using the of OR 2-4 for reduced penetrance for *BRCA1/BRCA2*, as this is the definition of moderate-penetrance used for genes such as *ATM* and *CHEK2,* for which VCEP specifications of the ACMG already exist or are under development^12^ ^27^.

### Classification of rare variants with and without evidence of effect size

All variants (and all VUS) have a true underlying effect size (association with disease): this may be of standard penetrance (OR≥4 in this use case), reduced penetrance (OR 2-4), marginal association (OR 1-2) or neutrality/protective (OR≤1). For variants of reasonable frequency, observed human data may be available by which this effect size is directly estimable (albeit confidence intervals may be wide). However, for the vast majority of newly-encountered variants (even in *BRCA1/BRCA2*), such data are not available and we can only infer this effect size from the summation of other available evidence types (only indirectly indicative of the underlying effect size). Across all the variants in a given gene (i) availability of this range of potential evidence items is highly incomplete and (ii) the correlation of evidence weightings with underlying effect size is rather crude for most evidence types. This is exemplified by inference of effect size from observation of biallelic variants in Fanconi anaemia, in which the phenotypic range is broad and at least one variant must cause incomplete loss of function to avoid embryonic lethality^28^.

An effect size of standard penetrance is inferred in the absence of direct quantitation of effect size if the weight of these other evidence items is deemed sufficient (routine ACMG/AMP variant interpretation approaches); in Scenario B an effect size of reduced penetrance is inferred on the basis of a modestly lesser weight (or concordance) of other evidence items. In the absence of availability for every clinically-encountered rare variant of well-powered case-control data, we shall continue to rely on such inferences if we seek to classify more than a tiny fraction of variants out of the VUS space. Nevertheless, even with implementation of this framework for Reduced penetrance, the majority of rare variants will remain as VUS due to a paucity of available data supporting pathogenicity. Of course, a subset of these “data-poor” VUS will have a true underlying effect size in the moderate or standard penetrance range; with a low-scoring *in silico* prediction, such a VUS could readily be classified as likely benign.

It may be possible that we can better refine variant-specific risks as larger case-control datasets become available. However, availability of larger population-control data adds little extra power unless there is concomitant availability of larger series from cases of the respective diseases. Furthermore, each case-control dataset will pertain to only one of potentially multiple associated phenotypes. Alternatively, and more plausibly, advances in high throughput functional assays may improve our estimates of variant-specific risk (although validation of this hypothesis would in itself require variant truth-sets accurately characterised for variant-specific effect sizes)^29^.

### Validation and future evolution of this Framework

It would have been optimal to evaluate the framework using a validated “truth-set” of variants validated as being of reduced penetrance (with each variant fully characterised for each evidence type). Such a “truth-set” would enable quantitative assessment for evidence allocation for different evidence items proposed within the Reduced Penetrance Framework. In practice, such a “truth-set” does not exist; our framework development was thus restricted to empirical adjustment of the existing ACMG framework via iterative expert consensus agreement.

To evaluate downstream real world clinical application of this framework within the CanVIG-UK community (and beyond), we have adapted our national variant platform (CanVar-UK) (i) to capture for a variant whether evaluation using the reduced penetrance framework was considered and (ii) to allow ‘Likely Pathogenic (Reduced Penetrance) as a formal classification option. Along with iterative real-time consultation regarding these variants through the CanVar-UK Diagnostic Users email forum, the new fields will enable formal review at one year of variants being evaluated as potentially of reduced penetrance.

## Limitations

Limitations of the framework development process include limited response rates for email survey send-outs as a proportion of CanVIG-UK membership and incomplete participation by attendees during in-meeting live polling. This may in part reflect that a proportion of the CanVIG-UK community (likely the clinical geneticists/genetic counsellors) do not directly undertake variant classification and thus did not feel confident in responding to questions involving detailed evidence allocation/variant classification. For the live polls, we could not align responses to professional role. The proportion of respondents from each centre also varied.

In summary, we present a preliminary framework for application to variants in high-penetrance genes of putative reduced penetrance, developed in consultation with the CanVIG-UK clinical network of diagnostic clinical scientists and genetics clinicians. The framework will be reviewed and updated following wider application in clinical practice. We anticipate that this framework will improve consistency in clinical approaches to classification of variants as being of reduced penetrance, nationally and beyond.

## Data Availability

All data are presented in the manuscript or supplementary materials.

## Ethics Declaration Statement

I confirm all relevant ethical guidelines have been followed. This work did not require IRB or ethics committee approvals. Only summary-level data from publicly available resources were used in this work.

## Author Contributions

Conceptualization: C.T., A.G.; Data curation: A.G., S.A.; Formal analysis: A.G., S.A.; Funding acquisition: C.T.; Investigation: A.G., S.A.; Methodology: C.T., A.G., S.A., C.F.R; Project administration: S.A.; Software: C.F.R.; Supervision: C.T.; Visualization: S.A., A.G.; Validation: M.D., G.J.B., A.C., J.F., B.F., S.P-S., J.G., T.McD., T.McV., H.H., C.F.R. Writing-original draft: C.T., A.G., S.A.; Writing-review & editing: all authors.

