## Supplementary Material for "Classification of Variants of Reduced Penetrance in High Penetrance Cancer Susceptibility Genes: Framework for Genetics Clinicians and Clinical Scientists by CanVIG-UK (Cancer Variant Interpretation Group-UK)"

### **Supplementary Methods**

The below process for the development of a framework for the classification of variants of reduced penetrance was followed:

1. Online survey on confidence in classifying variants with reduced penetrance and perceived utility of recommendation development circulated to CanVIG-UK members ahead of national CanVIG-UK meeting.
2. Hypothetical candidate reduced penetrance variants presented at national CanVIG-UK meeting with in-meeting polls on the application of evidence and overall classification (poll results for each question hidden until poll had closed). Verbal explanation of context provided with opportunity for clarification of questions prior to response.
3. Draft framework for classification of reduced penetrance variants developed by working group (CStAG). The context was divided into i) variants with quantified effect size (from case-control/segregation data) and ii) variants with inconsistent/weak evidence.
4. Draft framework for classification of reduced penetrance variants circulated to CanVIG-UK membership for comment and review ahead of national CanVIG-UK meeting.
5. Presentation of draft framework for classification of reduced penetrance variants with application of framework to exemplar variants at national CanVIG-UK meeting. In-meeting poll of application of evidence to exemplar variants and attitudes to draft framework (poll results for each question hidden until poll had closed). Verbal explanation of context provided with opportunity for clarification questions prior to response.
6. Modification of framework for classification of variants of reduced penetrance by working group (CStAG) based on feedback from CanVIG-UK.
7. Online survey on application of modified framework for classification of variants of reduced penetrance to hypothetical exemplar variants circulated to CanVIG-UK members ahead of national CanVIG-UK meeting. Participants had already been introduced to the framework at prior CanVIG-UK meetings and so were equipped to give more detailed responses in their own time.
8. Results of pre-meeting survey on application of modified draft framework to hypothetical exemplar variants discussed at national CanVIG-UK meeting with in-meeting poll of attitudes to modified framework (poll results for each question hidden until poll had closed).
9. Framework modified and finalised by working group (CStAG) following peer review and approved following circulation to CanVIG-UK group.

### **Supplementary Figures**

**Supplementary Figure 1: Additional results from scoping survey.** Questions were asked in the context of variant interpretation for cancer susceptibility genes.

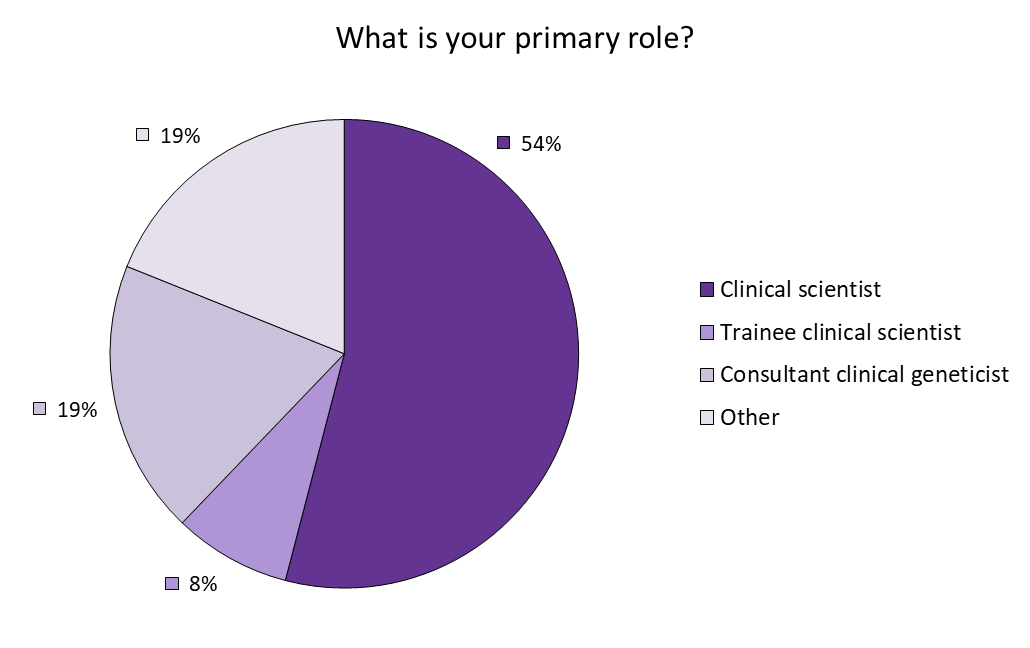

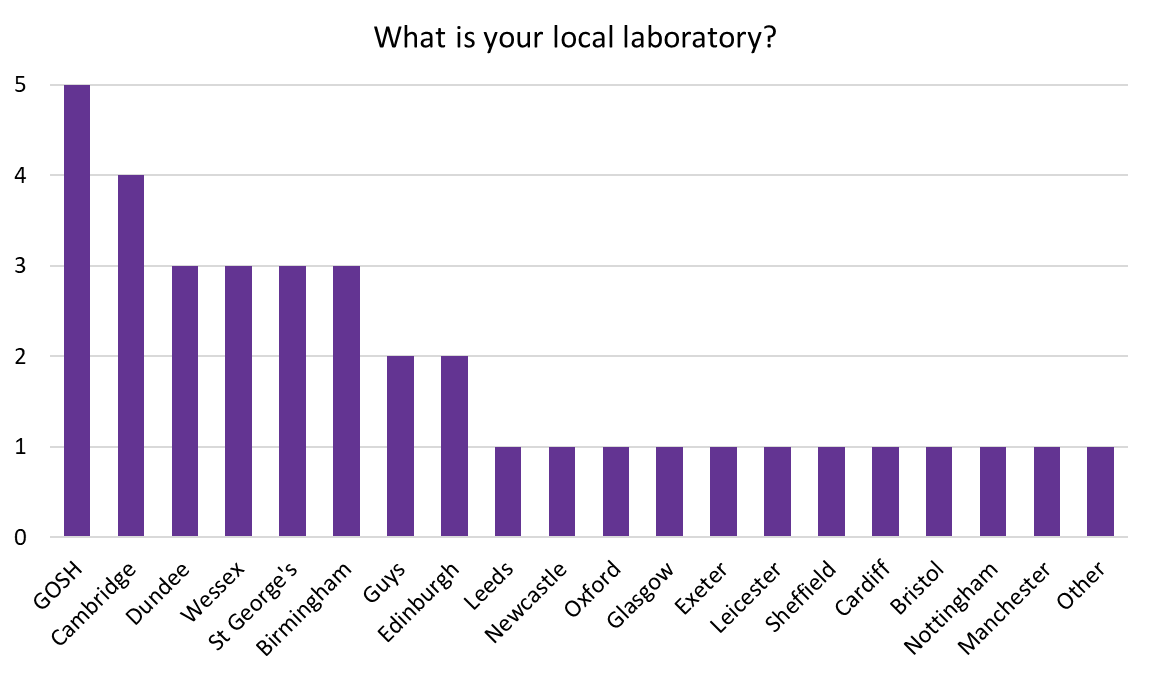

**A**

**B**

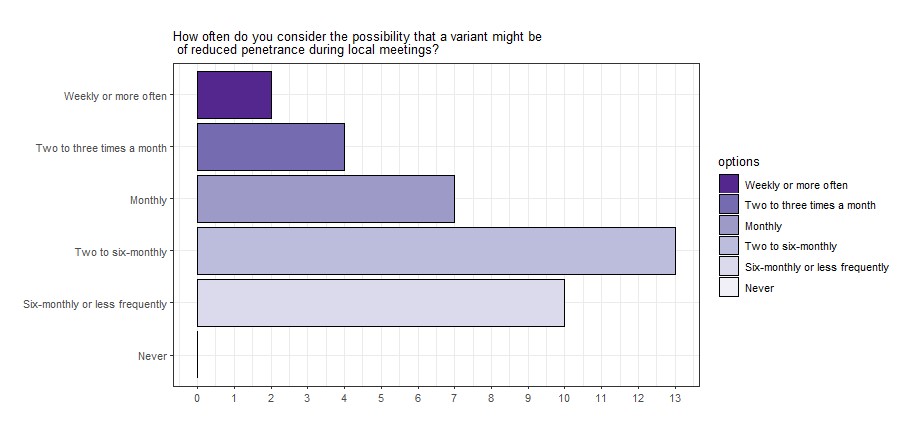

**C**

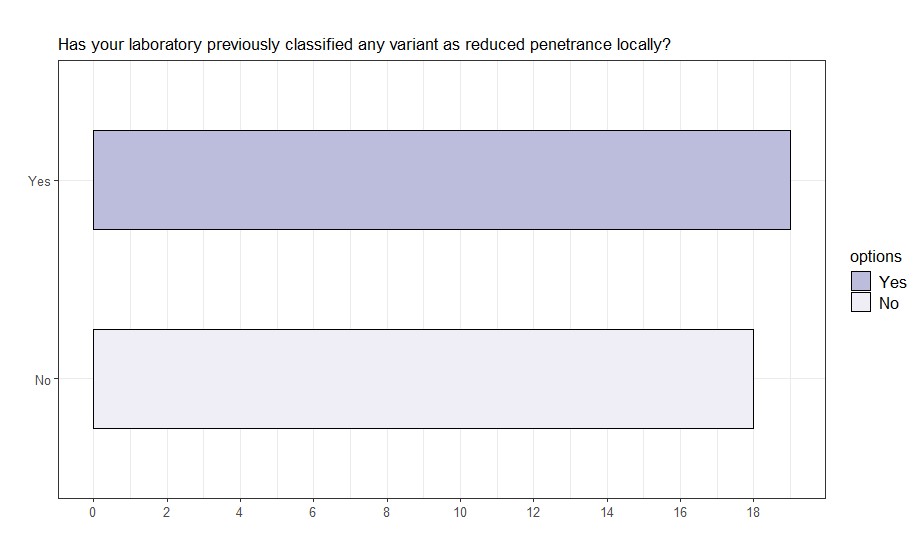

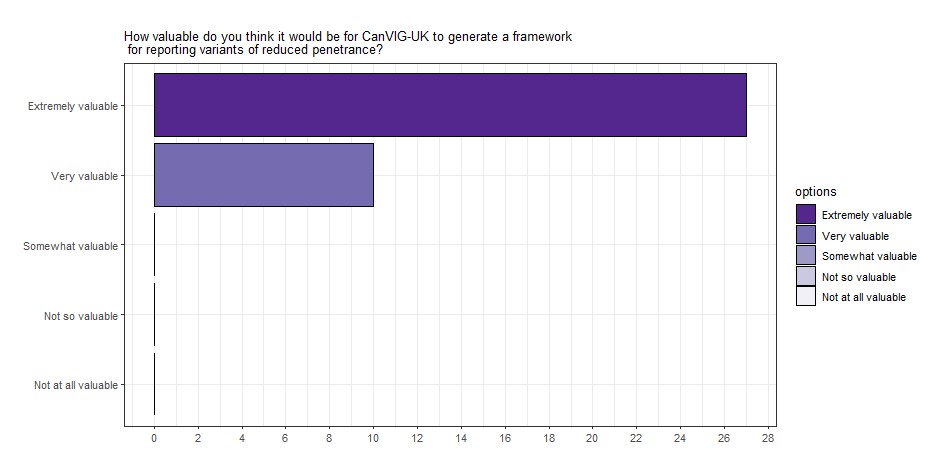

**D**

**E**

#### **Supplementary Figure 2: Draft consensus framework circulated prior to the February 2024 CanVIG-UK meeting**

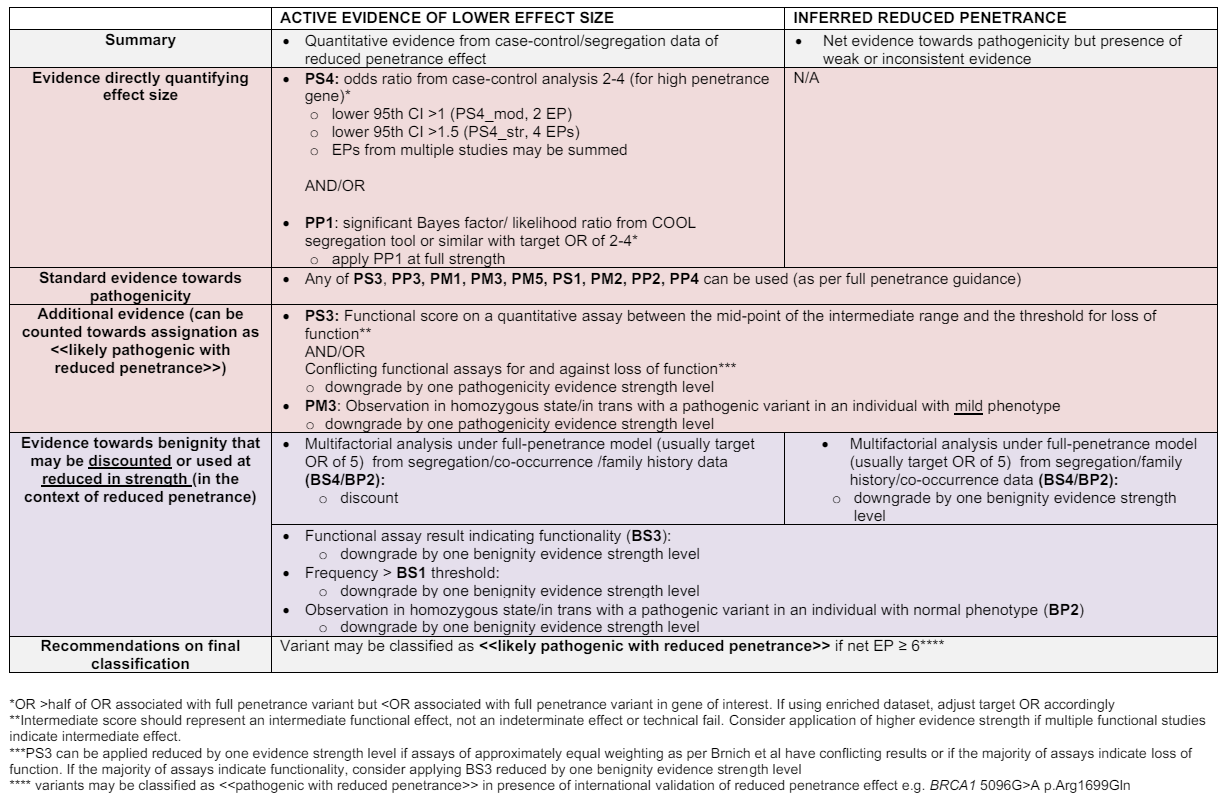

#### **Supplementary Figure 3: Live poll classification of ‘Scenario B’ variant NM_000059.4:c.8351G>A p.(Arg2784Gln)**

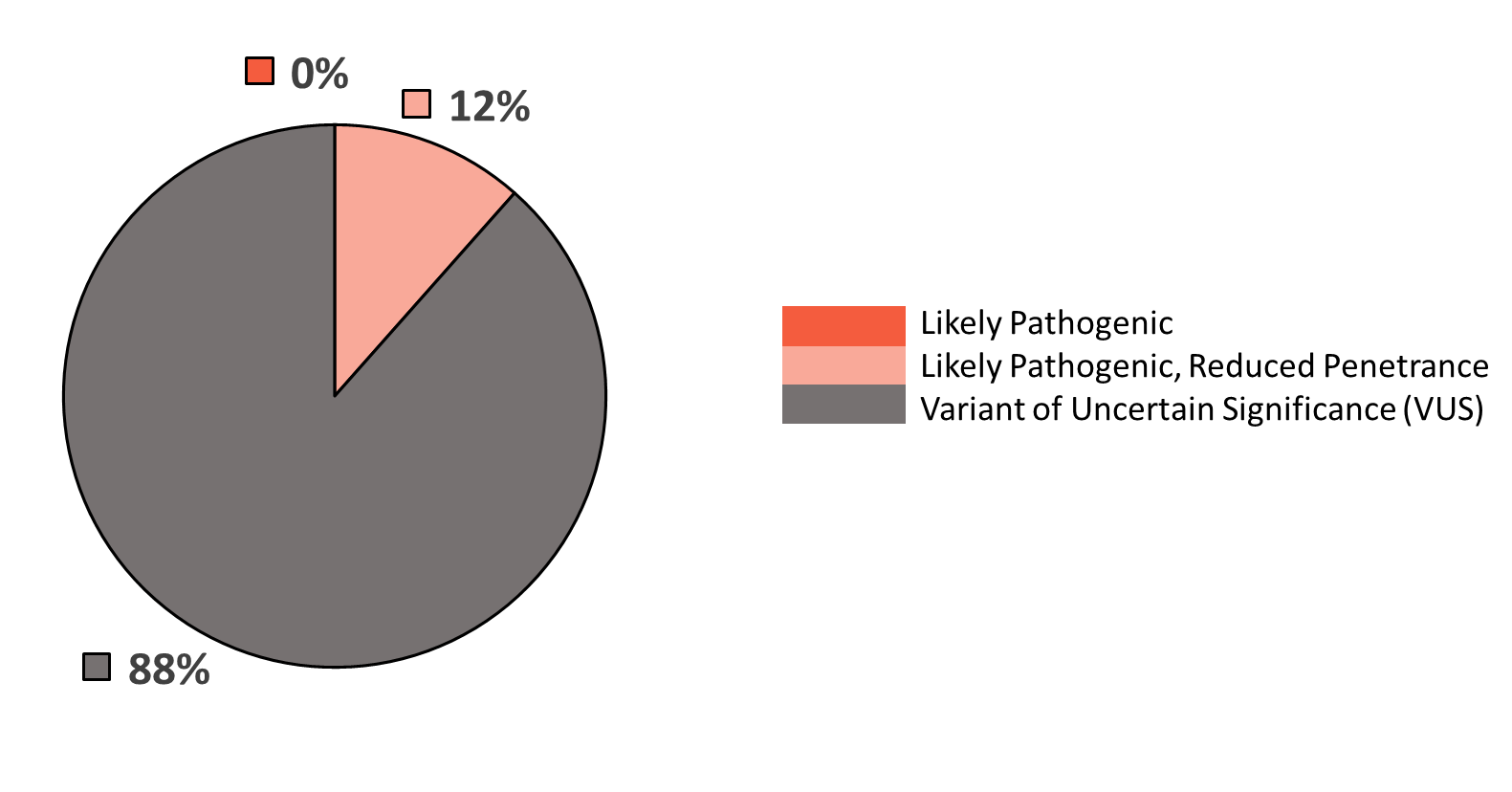

### **Supplementary Tables**

#### **Supplementary Table 1: Full results for all scenarios from the evidence allocation live poll (September 2023 CanVIG-UK meeting)**

|  | | | **Scoring of evidence item (reduced penetrance)** | | | | | | | | | | | **Overall classification of variant (reduced penetrance)** | | | | | **MDT discussion  required** | | |
| --- | --- | --- | --- | --- | --- | --- | --- | --- | --- | --- | --- | --- | --- | --- | --- | --- | --- | --- | --- | --- | --- |
|  |  |  | **Towards Benignity** | | | | | **Nil** | **Towards Pathogenicity** | | | | |  |  |  |  |  |  |  |  |
| **Poll Question** | **Evidence being scored** | **Other Evidence available for hypothetical variant** | Very Strong (-8 evidence points) | Strong (-4 evidence points) | -3 evidence points | Moderate (-2 evidence points) | Supporting (-1 evidence points) | Evidence not applied | Supporting (1 evidence point) | Moderate (2 evidence points) | 3 evidence points | Strong (4 evidence points) | Very Strong (8 evidence points) | Pathogenic | Likely Pathogenic | Variant of Uncertain Significance (VUS) | Likely Benign | Benign | None | Local | National |
| **Scenario 1:** How would you score this case control evidence towards pathogenicity (PS4)? | OR=1.5 (95% CI 1.0-2.0, p=0.05) | BRCT Domain REVEL = 0.76 1/118,479 observations in gnomAD v2.1.1 exomes | NA | NA | NA | NA | NA | 19 | 6 | 0 | 1 | 0 | 0 | 0 | 1 | 24 | 0 | 0 | 8 | 12 | 9 |
|  | OR=2.2 (95% CI 1.0-3.4, p=0.05) |  | NA | NA | NA | NA | NA | 6 | 6 | 11 | 0 | 0 | 0 | 0 | 4 | 20 | 0 | 0 | 7 | 8 | 8 |
|  | OR=2.2 (95% CI 1.5-2.7, p=0.001) |  | NA | NA | NA | NA | NA | 1 | 8 | 11 | 0 | 6 | 0 | 2 | 9 | 16 | 0 | 0 | 4 | 12 | 13 |
|  | OR=2.2 (95% CI 1.5-2.7, p=0.001)  and OR=2.2 (95% CI 1.0-3.4, p=0.05) |  | NA | NA | NA | NA | NA | 1 | 3 | 5 | 3 | 11 | 1 | 3 | 12 | 7 | 0 | 0 | 6 | 7 | 7 |
| **Scenario 2:** How would you score functional evidence towards benignity/pathogenicity (PS3/BS3)? *All assays have been assessed as providing 'strong' evidence on Brnich assessment using negative and positive control variants presumed as full penetrance.* | LOF on assay | BRCT domain  REVEL = 0.76  1/118,479 observations in gnomAD v2.1.1 exomes  OR=2.2 (95% CI 1.5-2.7, p=0.001) | 0 | 0 | 0 | 0 | 0 | 0 | 2 | 3 | 2 | 15 | 0 | 7 | 16 | 0 | 0 | 0 | 9 | 7 | 3 |
|  | Intermediate effect, towards LOF, on assay |  | 0 | 0 | 0 | 0 | 0 | 2 | 5 | 11 | 2 | 2 | 0 | 0 | 13 | 6 | 0 | 0 | 6 | 7 | 6 |
|  | Intermediate effect, towards functional |  | 0 | 0 | 1 | 1 | 1 | 10 | 5 | 1 | 0 | 1 | 0 | 0 | 3 | 16 | 0 | 0 | 5 | 5 | 5 |
|  | LOF on one assay,  Functional on different assay |  | 0 | 0 | 0 | 0 | 0 | 16 | 1 | 0 | 0 | 0 | 0 | 0 | 3 | 14 | 0 | 0 | 3 | 1 | 10 |
|  | Functional on assay |  | 0 | 13 | 0 | 4 | 2 | 1 | 0 | 1 | 0 | 0 | 0 | 0 | 2 | 13 | 4 | 0 | 7 | 6 | 3 |
| **Scenario 3:** How would you score this allelic evidence towards benignity/pathogenicity (BS2/PM3)? | Variant *in trans* with a BRCA1 pathogenic variant in individual with early onset breast cancer (no other phenotype). Abnormal chromosome breakage studies |  | 0 | 1 | 1 | 0 | 2 | 3 | 5 | 6 | 0 | 0 | 0 | 0 | 2 | 16 | 2 | 1 | 5 | 5 | 7 |
| **Scenario 4:** How would you score this allelic evidence towards benignity (BS2)? | Variant *in trans* with pathogenic BRCA2 variant, no reported Fanconi anaemia features. No available chromosome breakage studies (BS2) |  | 0 | 0 | 0 | 6 | 13 | 4 | NA | NA | NA | NA | NA | 1 | 3 | 19 | 2 | 0 | 4 | 7 | 10 |
| **Scenario 5:** How would you score this segregation evidence towards benignity (BS4)? *Segregation score generated under a full penetrance model*. | Segregation data from Parsons et al., 2019: LR = 0.04, -4 exponent points (BS4) |  | 0 | 4 | 2 | 8 | 4 | 4 | NA | NA | NA | NA | NA | 1 | 3 | 16 | 3 | 0 | 2 | 6 | 6 |

**Supplementary Table 2: Full responses from CanVIG-UK live polls regarding use of the reduced penetrance framework for ‘Scenario A’ and ‘Scenario B’ variants.** Live polls were conducted during the CanVIG-UK February and March 2024 meetings, with an online survey circulated between February and March meetings. Further information regarding the evidence discussed for each variant is provided in Table 2.

| **Poll/Survey** | **Question posed** | **Response options** | **Counts** |
| --- | --- | --- | --- |
| Live Poll (in-meeting, February 2024) | Using the reduced penetrance framework, how would you classify the following variant: *BRCA2* c.9302T>G p.(Leu3101Arg)? | Likely Pathogenic | 4 |
|  |  | Likely Pathogenic, reduced penetrance | 16 |
|  |  | Variant of Uncertain Significance | 1 |
|  | Using the reduced penetrance framework, how would you classify the following variant: *BRCA2* c.520C>T p.(Arg174Cys)? | Likely Pathogenic | 0 |
|  |  | Likely Pathogenic, reduced penetrance | 21 |
|  |  | Variant of Uncertain Significance | 5 |
|  | Using the reduced penetrance framework, how would you classify the following variant: *BRCA2* c.8351G>A p.(Arg2784Gln)? | Likely Pathogenic | 0 |
|  |  | Likely Pathogenic, reduced penetrance | 3 |
|  |  | Variant of Uncertain Significance | 23 |
|  | I would be comfortable as classifying a variant as likely pathogenic with reduced penetrance if supported by case-control evidence | Agree | 25 |
|  |  | Disagree | 2 |
|  | I would be comfortable as classifying a variant as likely pathogenic with reduced penetrance if supported by segregation evidence | Agree | 21 |
|  |  | Disagree | 6 |
|  | I would be comfortable as classifying a variant as likely pathogenic with reduced penetrance in absence of case/control or segregation effect size | Agree | 7 |
|  |  | Disagree | 19 |
| Online Survey (between February and March 2024 CanVIG-UK meetings) | At what evidence strength would you apply evidence towards benignity (BS2) for variant (i)? | None (would not apply) | 2 |
|  |  | BS2 supporting | 2 |
|  |  | BS2 moderate | 7 |
|  |  | BS2 strong | 0 |
|  | Using the reduced penetrance framework, how would you classify variant (i)? | Likely Pathogenic | 4 |
|  |  | Likely Pathogenic, reduced penetrance | 6 |
|  |  | Variant of Uncertain Significance | 1 |
|  | At what evidence strength would you apply evidence towards benignity (BP5) for variant (ii)? | None (would not apply) | 0 |
|  |  | BP5 supporting | 2 |
|  |  | BP5 moderate | 6 |
|  |  | BP5 strong | 2 |
|  | Using the reduced penetrance framework, how would you classify a variant such as hypothetical variant (ii)? | Likely Pathogenic | 1 |
|  |  | Likely Pathogenic, reduced penetrance | 2 |
|  |  | Variant of Uncertain Significance | 7 |
| Live Poll (in-meeting, March 2024) | Using the reduced penetrance framework, how would you classify a variant such as hypothetical variant (iii)? | Likely Pathogenic | 3 |
|  |  | Likely Pathogenic, reduced penetrance | 16 |
|  |  | Variant of Uncertain Significance | 2 |
|  | I would be comfortable as classifying a variant as likely pathogenic with reduced penetrance in absence of case/control or segregation effect size | Agree | 15 |
|  |  | Disagree | 9 |

### **Supplementary Note 1: CanVIG-UK Consortium Group Membership**

C. Turnbull^1,49^, A. Garrett^1^, L. Loong^1^, S. Choi^1^, B. Torr^1^, S. Allen^1^, M. Durkie^2^, A. Callaway^3^, J. Drummond^4^, G.J. Burghel^5^, R. Robinson^6^, I.R. Berry^65^, A.J. Wallace^5^, D.M. Eccles^7, 8^, M. Tischkowitz^13^, S. Ellard^9^, H. Hanson^1,16^, E. Baple^10,11^, D.G. Evans^5,30^, E. Woodward^5,30^, F. Lalloo^5,30^, S. Samant^33^, A. Lucassen^57,14,15^, A. Znaczko^44^, A. Shaw^23^, A. Ansari^34^, A. Kumar^21^, A. Donaldson^53^, A. Murray^19^, A. Ross^18^, A. Taylor-Beadling^22^, A. Taylor^18^, A. Innes^25^, A. Brady^29^, A. Kulkarni^23^, A.C. Hogg^5,^ A. Ramsay Bowden^18^, A. Hadonou^47^, B. Coad^16^, B. McIldowie^19^, B. Speight^18^, B. DeSouza^47^, B. Mullaney^3^, C. McKenna^62^, C. Brewer^44^, C. Olimpio^18^, C. Clabby^40^, C. Crosby^47^, C. Jenkins^42^, C. Armstrong^33^, C. Bowles^9^, C. Brooks^22^, C. Byrne^62^, C. Maurer^4^, D. Baralle^57^, D. Chubb^1^, D. Stobo^34^, D. Moore^35^, D.O'Sullivan^33^, D. Donnelly^62^, D. Randhawa^24^, D. Halliday^41^ , E. Atkinson^50^, E. Rauter^24^, E. Johnston^38^, E. Maher^8^, E. Sofianopoulou^17^, E. Petrides^23^, F. McRonald^43^, F. Pelz^51^, I. Frayling^19^, G. Corbett^62^, G. Rea^62^, H. Clouston^5^, H. Powell^31^, H. Williamson^52^, H. Carley^47^, H.J.W. Thomas^26^, I. Tomlinson^63^, J. Cook^46^, J. Hoyle^21^, J. Tellez^32^, J. Whitworth^18^, J. Williams^49^, J. Murray^35^, J. Campbell^27^, J. Tolmie^33^, J. Field^38^, J. Mason^64^, J. Burn^31^, J. Bruty^18^, J. Callaway^8^, J. Grant^34^, J. Del Rey Jimenez^47^, J. Pagan^35^, J. VanCampen^24^, J. Barwell^53^, K. Monahan^29^, K. Tatton-Brown^16,^ K.R. Ong^63^, K. Murphy^33^, K. Andrews^18,^ K. Mokretar^23^, K. Cadoo^48^, K. Smith^52^, K. Baker^8^, K. Brown^24^, K. Reay^64^, K. McKay Bounford^34^, K. Bradshaw^38^, K. Russell^65^, K. Stone^23^, K. Snape^16^, L. Crookes^5^, L. Reed^21^, L. Taggart^62^, L. Yarram^65^, L. Cobbold^47^, L. Walker^39^, L. Walker^41^, L. Hawkes^16^, L. Busby^22^, L. Izatt^23^, L. Kiely^22,^ L. Hughes^64^, L. Side^56^, L. Sarkies^18^, K.-L. Greenhalgh^28^, M. Shanmugasundaram^63^, M. Duff^40^, M. Bartlett^29^, M. Watson^3^, M. Owens^9^, M. Bradford^54^, M. Huxley^64^, M. Slean^33^, M. Ryten^23^, M. Smith^55^, M. Ahmed^21^, N. Roberts^2^, C. O'Brien^50^, O. Middleton^33^, P. Tarpey^4^, P. Logan^62^, P. Dean^3^, P. May^24^, P. Brace^21^, R. Tredwell^38^, R. Harrison^37^, R. Hart^63^, R. Kirk^5^, R. Martin^31^, R. Nyanhete^3^, R. Wright^2^, R. Martin^62^, R. Davidson^34^, R. Cleaver^45^, S. Talukdar^16^, S. Butler^64^, J. Sampson^19^, S. Ribeiro^49^, S. Dell^46^, S. Mackenzie^32^, S. Hegarty^62^, S. Albaba^5^, S. McKee^36^, S. Palmer-Smith^19^, S. Heggarty^62^, S. MacParland^62^, S. Greville-Heygate^49^, S. Daniels^4^,S. Prapa^18^,S. Abbs^4^, S. Tennant^33^, S. Hardy^43^, S. MacMahon^49^, T. McVeigh^49^, T. Foo^49^, T. Bedenham^42^, T. Cranston^42,^ T. McDevitt^40^, V. Clowes^29^, V. Tripathi^23^, V. McConnell^62^, N. Woodwaer^45^, Y. Wallis^64^, Z. Kemp^49^, G. Mullan^62^, L. Pierson^62^, L. Rainey^62^, C. Joyce^59^, A. Timbs^41^, A-M. Reuther^3^, B. Frugtniet^47^, B. DeSouza^25^, C. Husher^3^, C. Lawn^22^, C. Corbett^63^, D. Nocera-Jijon^16^, D. Reay^31^, E. Cross^3^, F. Ryan^3^, H. Lindsay^6^, J. Oliver^6^, J. Dring^63^, J. Spiers^65^, J. Harper^23^, K. Ciucias^34^, L. Connolly^60^, M. Tsang^62^, R. Brown^6^, S. Shepherd^32^, S. Begum^16^, S. Daniels^3^, T. Tadiso^16^, T. Linton-Willoughby^4^, H. Heppell^35^, K. Sahan^61^, L. Worrillow^6^, Z. Allen^22^, C. Watt^34^,M. Hegarty^62^, R. Mitchell^6^, R. Coles^66^, G. Nickless^23^, E. Cojocaru^49^, I. Doal^64^, F. Sava^64^, C. McCarthy^62^, R. Jeeneea^63^, D. Goudie^20^, M. McConachie^20^, S. Botosneanu^5^, G. Kavanaugh^1^, K. Russell^10^, C. Sherlaw^63^, O. Tsoulaki^46^, C. Forde^5^, E. Petley^63^, A-B. Jones^1^, K. Oprych^16^, S. Pryde^67^, Z. Hyder^5^, N. Elkhateeb^18^, R. Braham^21^, L. Hanington^41^, C. Huntley^1^, R. Irving^51^, A. Sadan^23^, M. Ramos^22^, C. Elliot^35^, D. Wren^22^, D.Lobo^35^, J. McLean^68^, D. May^18^, L. Kearney^48^, T. Campbell^38^, K. Asakura^68^, L. Alwadi^19^, R. O’Shea^48^, J. Gabriel^42^, L. Chiecchio^3^, P. Bowman^44^, L.A. Sutton^48^, C. Walsh^23^, V. Cloke^69^, D. Ucanok^37^, J. Davies^65^, B. Pleasance^65^, E. Maguire^6^, A. Whaite^70^, S. Best^71^, S. Westbury^72^, A. Logan^62^, D. Navarajasegaran^71^, A. Bench^35^, P. Wightman^34^, A. Cartwright^2^, E. Higgs^41^, J.Bott^42^, H. Whitehouse^5^, J. Stevens^58^, D. Martin^37^, J. Dunlop^68^, S. Thomas^73^, C. Sau^68^, S. Farndon^74^, N. Coleman^48^, P. Angelini^49^, M. Duff^59^, H. Massey^35^, C. Rowlands^1^, C. Garcia-Petit^68^, K. Gillespie^68^, A. Alder^68^, E. Middleton^68^, C. Cassidy^75^, N. Orfali^48^, A. Webb^3^, A. Luharia^64^, N. Walker^33^, J. Charlton^71^, A. Andreou^47^, J. Peddie^68^, M. Khan^23^, L. Wilkinson^31^, H. Bezuidenhout^47^, M. Edis^18^, A. Callard^29^, P. Ostrowski^76^, P. Moverley^51^, K. Bean^73^, A. Dunne^48^, A. Moleirinho^23^, S. Waller^5^, K. Cox^47^, L. Greensmith^28^, A. Brittle^5^, N. Gossan^5^, L. Freestone^4^, C. Shak^77^, T. Langford^75^, Y. Clinch^21^, H. Livesey^51^, S. Borland^46^, A. Joshi^47^, K. Wall^77^, A. Whitworth^46^, A.Wilsdon^37^, K. Edgerley^72^, S. Pugh^5^, N. Chrysochoidi^3^, S. Mutch^38^, C. McMullan^5^, Y. Johnston^78^, M. Muraru^77^, A. May^77^, R. Begum^77^, C. Smith^44^, R. Patel^47^, I. Bhatnagar^79^, A. Taylor^62^, D. Brown^69^, J. Willan^46^, S. Taylor^48^, K. Jones^21^, K. Cox^21^, C. Ramsden^75^, O. Taiwo^49^, J. Jaudzemaite^47^, R. Sharmin^47^, L. Young^34^, C.O’Dubhshlaine^35^, L. McSorley^80^, I. Abreu Rodriguez^49^, S. Lillis^23^, P. Alexopoulos^23^, E. Mortensson^65^, L. Kingham^4^, R. Moore^18^, M. Kosicka-Slawinska^81^, S. Aslam^65^, R. Wells^82^, A. Carter^82^, H. Warren^6^, E. Rolf^49^, H. Reed^75^, L. Pearce^83^, D. Lock^6^, F. Ali^66^, A. Kolozi^84^, N. White^48^, D. Wood^34^, C. Hayden^25^

^1^ Division of Genetics and Epidemiology, Institute of Cancer Research, Sutton, UK

^2^ Yorkshire and North East Genomic Laboratory Hub, Sheffield Children's NHS Foundation Trust, Sheffield, UK

^3^ Central and South Genomics Laboratory Hub, Wessex Genomics Laboratory Service, Salisbury NHS Foundation Trust, Salisbury, UK

^4^ East Genomic Laboratory Hub, Cambridge University Hospitals Genomic Laboratory, Cambridge University NHS Foundation Trust, Cambridge, UK

^5^ Manchester Centre for Genomic Medicine and North West Genomic Laboratory Hub, Manchester University NHS Foundation Trust, Manchester, UK

^6^ Yorkshire and North East Genomic Laboratory Hub, Leeds Teaching Hospitals NHS Trust, Leeds, UK

^7^ Cancer Sciences, Faculty of Medicine, University of Southampton, Southampton, UK

^8^ Human Genetics and Genomic Medicine, Faculty of Medicine, University of Southampton, Southampton, UK

^9^ Department of Molecular Genetics, Royal Devon and Exeter NHS Foundation Trust, Exeter, UK

^10^ Genomics England, London, UK

^11^ University of Exeter Medical School, Exeter, UK

^12^ Division of Evolution & Genomic Sciences, The University of Manchester

^13^ Department of Medical Genetics, National Institute for Health, Research Cambridge Biomedical Research Centre, University of Cambridge, Cambridge, UK

^14^ Wessex Clinical Genetics Service, University Hospital Southampton NHS Foundation Trust, Southampton, UK

^15^ Clinical Ethics and Law Unit, University of Southampton, Southampton, UK

^16^ Department of Clinical Genetics, St. George's University Hospitals NHS Foundation Trust, London, UK

^17^ Public Health and Primary Care, Clinical Medicine, University of Cambridge, Cambridge, UK

^18^ Cambridge University Hospitals NHS Foundation Trust, Cambridge, UK

^19^ Institute of Medical Genetics, University Hospital of Wales, Cardiff and Vale University Health Board, Cardiff, UK

^20^ East of Scotland Regional Genetics Service, Level 6, Ninewells Hospital, Dundee

^21^ Great Ormond Street Hospital for Children NHS Foundation Trust, London, UK

^22^ North Thames Genomic Laboratory Hub, Great Ormond Street Hospital for Children NHS Foundation Trust, London, UK

^23^ Department of Clinical Genetics, Guy’s and St Thomas’ NHS Foundation Trust, London, UK

^24^ South East Genomic Laboratory Hub, Guy’s and St Thomas’ NHS Foundation Trust, London, UK

^25^ Genomic Medicine Service, Imperial College Healthcare NHS Trust, London, UK

^26^ Faculty of Medicine, Department of Surgery & Cancer, Imperial College London, London, UK

^27^ Institute of Neurology, UCL Queen Square Institute of Neurology, London, UK

^28^ Liverpool Women’s NHS Foundation Trust, Liverpool, UK

^29^ London North West University Healthcare NHS Trust, London, UK

^30^ Division of Evolution and Genomic Sciences, School of Biological Sciences, Faculty of Biology Medicine and Health, The University of Manchester, Manchester, UK.

^31^ The Newcastle upon Tyne Hospitals NHS Foundation Trust, Newcastle upon Tyne, UK

^32^ North East and Yorkshire Genomic Laboratory Hub, The Newcastle upon Tyne Hospitals NHS Foundation Trust, Newcastle upon Tyne, UK

^33^ NHS Grampian, Aberdeen, UK

^34^ NHS Greater Glasgow and Clyde, Glasgow, UK

^35^ NHS Lothian, Edinburgh, UK

^36^ Northern Ireland Regional Genetics Service, Belfast Health & Social Care Trust, Belfast, UK

^37^ Nottingham University Hospitals NHS Trust, Nottingham, UK

^38^ East Midlands and East of England Genomics Laboratory, Nottingham University Hospitals NHS Trust, Nottingham, UK

^39^ University of Otago, Otago, New Zealand

^40^ Our Lady's Children's Hospital, Crumlin, Dublin, Ireland

^41^ Clinical Genetics, Oxford University Hospitals NHS Foundation Trust, Oxford, UK

^42^ West Midlands, Oxford and Wessex Genomic Laboratory Hub, Oxford University Hospitals NHS Foundation Trust, Oxford, UK

^43^ Public Health England, London, UK

^44^ Royal Devon and Exeter NHS Foundation Trust, Exeter, UK

^45^ Royal Free London NHS Foundation Trust, London, UK

^46^ Sheffield Children's NHS Foundation Trust, Sheffield, UK

^47^ St George’s University Hospitals NHS Foundation Trust, London, UK

^48^ St James’s Hospital, Dublin, Ireland

^49^ Cancer Genetics Unit, The Royal Marsden NHS Foundation Trust, Sutton, London, UK

^50^ Trinity College Dublin, The University of Dublin, Ireland

^51^ University Hospital of Wales, Cardiff and Vale University Health Board, Cardiff, UK

^52^ University Hospitals Bristol NHS Foundation Trust, Bristol, UK

^53^ University Hospitals of Leicester NHS Trust, Leicester, UK

^54^ University Hospitals of Plymouth NHS Trust, Plymouth, UK

^55^ University of Manchester, Manchester, UK

^56^ Wessex Clinical Genetics Service, Princess Anne Hospital, Southampton, UK

^57^ Faculty of Medicine, University of Southampton, Southampton, UK

^58^ University Hospital Southampton NHS Foundation Trust, Southampton, UK

^59^ Cork University Hospital, Cork, Ireland

^60^ Children’s Health Ireland (CHI), Crumlin, Dublin, Ireland

^61^ The Ethox Centre, Oxford, UK

^62^ Belfast Health & Social Care Trust, Belfast, UK

^63^ Birmingham Women’s and Children’s NHS Foundation Trust, Birmingham, UK

^64^ Central and South Genomic Laboratory Hub, Birmingham Women’s and Children’s NHS Foundation Trust, Birmingham, UK

^65^ Bristol Genetics Laboratory, Pathology Sciences, Southmead Hospital, North Bristol NHS Trust, Bristol, United Kingdom

^66^ Northwick Park Hospital, Watford Rd, Harrow, UK

^67^ Chapel Allerton Hospital, Chapeltown Rd, Leeds, UK

^68^ NHS Tayside, UK

^69^ South East Scotland Genetic Service, Western General Hospital, Edinburgh, UK

^70^ Liverpool Centre for Genomic Medicine, Liverpool Women’s NHS Foundation Trust, Liverpool, UK

^71^ King’s College Hospital, London, UK

^72^ University Hospital Bristol and Weston NHS Foundation Trust, Bristol, UK

^73^ Sheffield Teaching Hospital NHS Foundation Trust, Sheffield, UK

^74^ Bristol Royal Hospital for Children, Bristol. UK

^75^ Manchester University Foundation Trust, Manchester, UK

^76^ North East Thames Clinical Genetics Service, London, UK

^77^ West Midlands Regional Genetics Laboratory, Birmingham Women’s Hospital, Birmingham, UK

^78^West of Scotland Centre for Genomic Medicine, Queen Elizabeth University Hospital, Glasgow, UK

^79^Oxford Centre for Genomic Medicine, Oxford University Hospitals NHS Foundation Trust, Oxford, UK

^80^St Vincent’s Hospital Group, Elm Park, Dublin, Ireland

^81^North West Thames Regional Genetics Service, St. Mark's Hospital, Harrow, UK

^82^Royal Liverpool University Hospital Trust, Liverpool, UK

^83^All Wales Medical Genetics Service, Cardiff, Wales, UK

^84^GenQA

### **Supplementary Note 2: CStAG Membership**

A. Garrett^1,2^, S. Allen^1^_,_ M Durkie^3^, G.J. Burghel^4^, R. Robinson^5^, A. Callaway^6^, J. Field^7^, B. Frugtniet^2^, S. Palmer-Smith^8^, J. Grant^9^, J. Pagan^10^, T. McDevitt^11^, T. McVeigh^12^, H. Hanson^1,13,14^, C. Turnbull^1,12^

1) Division of Genetics and Epidemiology, The Institute of Cancer Research, London, UK.
2) St George's University Hospitals NHS Foundation Trust, Tooting, London, UK
3) Sheffield Diagnostic Genetics Service, NEY Genomic Laboratory Hub, Sheffield Children's NHS Foundation Trust, Sheffield, UK
4) Manchester Centre for Genomic Medicine and NW Laboratory Genetics Hub, Manchester University Hospitals NHS Foundation Trust, Manchester, UK
5) The Leeds Genetics Laboratory, NEY Genomic Laboratory Hub, Leeds Teaching Hospitals NHS Trust, Leeds, UK
6) Central and South Genomics Laboratory Hub, Wessex Genomics Laboratory Service, University Hospital Southampton NHS Foundation Trust, Salisbury, UK.
7) Genomics and Molecular Medicine Service, Nottingham University Hospitals NHS Trust, Nottingham, UK
8) Institute of Medical Genetics, University Hospital of Wales, Cardiff and Vale University Health Board, Cardiff, UK
9) Laboratory Genetics, Queen Elizabeth University Hospital, NHS Greater Glasgow and Clyde, Glasgow, UK
10) South East Scotland Clinical Genetics, Western General Hospital, Edinburgh, UK.
11) Department of Clinical Genetics, CHI at Crumlin, Dublin, Ireland
12) The Royal Marsden NHS Foundation Trust, Fulham Road, London
13) Peninsula Regional Genetics Service, Royal Devon University Healthcare NHS Foundation Trust, Exeter, UK
14) Faculty of Health and Life Sciences, University of Exeter, Exeter, UK
